# Characterizing rurality using the *All of Us* research program data

**DOI:** 10.1101/2025.07.11.25331347

**Authors:** Michael Bradfield, Toluwanimi Olorunnisola, Vignesh Subbian

**Affiliations:** Banner Health North Colorado Medical Center, Department of Family Medicine, Greeley, Colorado, United States of America; College of Engineering, The University of Arizona, Tucson, Arizona, United States of America

**Keywords:** *All of Us* Research Program, Rural Health, Geolocation, Three-digit ZIP code

## Abstract

Rural communities experience disproportionately higher rates of chronic diseases, less access to healthcare services, and poorer health outcomes compared to their urban counterparts in the United States. However, inconsistencies in how rurality is defined across biomedical research, including limitations in geographic detail within large-scale datasets, present significant challenges for reliably studying rural health outcomes. This study aimed to develop and apply a scalable, operational rurality scale using 3-digit ZIP codes to characterize rural participation in the *All of Us* Research Program and to examine associations between rurality, delayed care, and healthcare affordability. Publicly available information from the Federal Office of Rural Health Policy and the Environmental Systems Research Institute was integrated to generate a continuous rurality scale at the 3-digit ZIP code level. A Kolmogorov-Smirnov test identified statistically significant differences in the geographic distribution of those who had delayed access to care (P < 0.001) and those with difficulties affording care (P < 0.001). The proposed continuous rurality scale is reproducible and extensible in several ways within the *All of Us* Workbench, as it provides a framework for categorizing participants by geolocation and facilitates standardized analyses of rurality-related research questions.

## Introduction

Rural communities experience disproportionately higher rates of chronic diseases, less access to healthcare services, and poorer health outcomes compared to their urban counterparts in the U.S. (1). Studying rural health is challenging, in part, due to the many definitions of “rurality” among researchers, healthcare entities, and the federal government. The notion of rurality encompasses many ideological, demographic, economic, and cultural constructs, though most acknowledge the definition should include some geographic component (2–5). Definitions and geographic units used to determine rurality vary widely across biomedical and health services research. These inconsistencies influence not only research findings but also policy and how health disparities are identified and resources allocated (6).

Depending on the source, approximately one in five Americans lives in rural areas (3,4,9). Rural health is adversely affected by many factors, including geographic isolation, inadequate healthcare infrastructure, physician shortages, poverty, low educational attainment, poor health literacy, and inadequate public transportation (2,10,11). Overall, this culminates in an elevated disease burden and decreased life expectancy for rural populations (11). One of the first steps in addressing these discrepancies is a fully operational and contextually relevant measure of rurality (8). This is especially important for research using large-scale, real-world health datasets such as the *All of Us* Research dataset (12), where geographic identifiers are typically restricted to 3-digit ZIP codes to protect participant privacy. Without a consistent and privacy-preserving method for defining rurality, efforts to examine rural health disparities and link geographic context to health outcomes remain limited in scope and impact.

The National Institute of Health’s *All of Us* Research Program is a precision medicine initiative that aims to enroll one million or more American participants (12,13). One of the primary goals of the program is to engage with and reduce health disparities among traditionally underserved groups, including those who are geographically underserved in biomedical research (13). The *All of Us* data holds significant value when studying rural health outcomes, as it includes a more diverse range of data types and sources than most existing data sets, including demographic data, geolocation data, survey responses, electronic health record (EHR) data, and genomic data (14).

The *All of Us* Research Program specifies that residents of established rural and non-metropolitan ZIP codes that meet the Health Resource and Services Administration (HRSA) Federal Office of Rural Health Policy (FORHP) rural grant eligibility criteria are underrepresented in biomedical research (15–17). The FORHP definition is broad and includes all non-metropolitan counties, certain commuting areas, and low-population density areas (15). Despite adopting the FORHP definition, the *All of Us* program does not provide any readily available method or indicator to identify and classify participants based on rurality within the data set.

Therefore, the purpose of this study is to develop and apply a rurality scale based on 3-digit ZIP codes to identify and characterize rural participation and enrollment within the *All of Us* Research Program. We then apply this scale to examine patterns in healthcare access and utilization, with the goal of informing future research on rural health disparities and enhancing the utility of large-scale datasets for rural health equity research.

## Methods

### *All of Us* program methods for recruitment and data collection

The *All of Us* Research Program recruits participants through multiple mechanisms, including academic medical centers, healthcare provider organizations, community-based enrollment sites, and digital platforms. Upon enrollment, participants complete a comprehensive informed consent process, which includes consent for long-term participation, sharing of electronic health records (EHRs), completion of health-related surveys, biospecimen donation (blood, saliva, and urine), and return of research results. The consent process also includes education about data use, privacy protections, and the ability to withdraw at any time (18,19). Participant data are then harmonized and standardized using the Observational Medical Outcomes Partnership (OMOP) Common Data Model (20). To protect participant privacy, *All of Us* data undergo a series of transformations, including removal of personally identifiable information, before being made available to researchers in a secure, cloud-based Researcher Workbench (21). This study was conducted under an approved Data Use Agreement and adhered to all ethical research conduct and data use policies established by the *All of Us* Research Program.

### *All of Us* researcher workbench

The *All of Us* Researcher Workbench operates under a data passport model, where authorized users can access data and execute research projects. The Workbench offers two primary tiers of data access that investigators can use for research purposes: a Registered Tier and a Controlled Tier. The Registered Tier consists of data from EHR, wearable devices, survey responses, and physical measurements. The Controlled Tier holds genomic data, including whole genome sequencing, genotyping arrays, and the first three-digit ZIP code geolocation data. The Workbench provides collaborative workspaces, an interactive Jupyter Notebook environment with the ability to perform analyses using R or Python, and tools for developing study cohorts. We performed our analysis using Python 3 (version 3.10.12) and utilized the *All of Us* Controlled Tier workspace using the version 8 curated dataset released in February 2025.

### Creating a rurality scale using geolocation codes

The *All of Us* program provides three-digit ZIP codes as part of the Controlled Tier dataset (22), in accordance with the Health Insurance Portability and Accountability Act (HIPAA) privacy rule for de-identifying Protected Health Information (23). However, restricting geographic detail to three-digit ZIP codes presents a methodological challenge for studying rural populations. To address this challenge, we constructed a rurality measure by analyzing two publicly available data sources: the Federal Office of Rural Health Policy (FORHP) dataset (15), which lists all rural U.S. ZIP codes and the Environmental Systems Research Institute (ESRI) dataset (24), which includes all ZIP codes nationally. We aggregated this information to generate a classification scheme for rural 3-digit ZIP codes, as illustrated in Fig 1.

**Fig 1.**
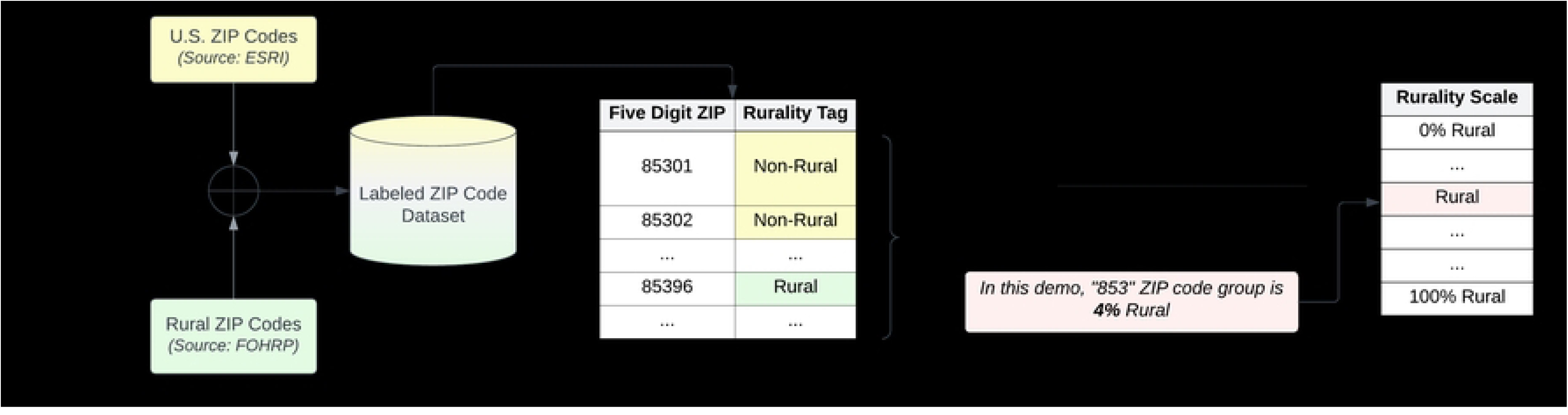
Demonstration of developing a Rurality Scale to study the rural-urban continuum when working with three-digit ZIP codes in a de-identified safe harbor dataset.

First, we obtained all U.S. five-digit ZIP codes and their corresponding population from the U.S. ESRI ZIP code geodatabase. The list of rural five-digit ZIP codes from the FORHP dataset was then used to tag all ZIP codes as “rural” or “non-rural.” Next, we grouped all five-digit ZIP codes based on their first three digits. For each three-digit ZIP code group, a rural percentage was computed by taking the ratio of the population of those codes marked as “rural” divided by the total population of that three-digit ZIP code group (Fig 1). Finally, each *All of Us* participant was mapped to a place in the rurality scale based on their corresponding three-digit ZIP code to enable further analysis.

### Healthcare access and utilization survey data preparation and analysis

In addition to geolocation codes (3-digit ZIP codes), we used the following data: demographics (age, sex, ethnicity, and race), educational status, and responses to the Healthcare Access and Utilization survey (25), which includes 114 questions (26). We reviewed all 114 questions and extracted those questions relevant to delayed care (9 questions) and healthcare affordability (14 questions). Delayed care was assessed using nine survey items related to healthcare access; participants with six or more affirmative responses were coded as 1, indicating significant experiences with delayed care, and fewer than six were coded as 0. Healthcare affordability was evaluated using fourteen items, with nine or more affirmative responses coded as 1, and fewer than nine coded as 0. Thresholds were selected to reflect a high burden of barriers in each domain and to capture participants with persistent or widespread challenges rather than isolated instances. Fig 2 illustrates the coding process for classifying participants’ responses.

**Fig 2.**
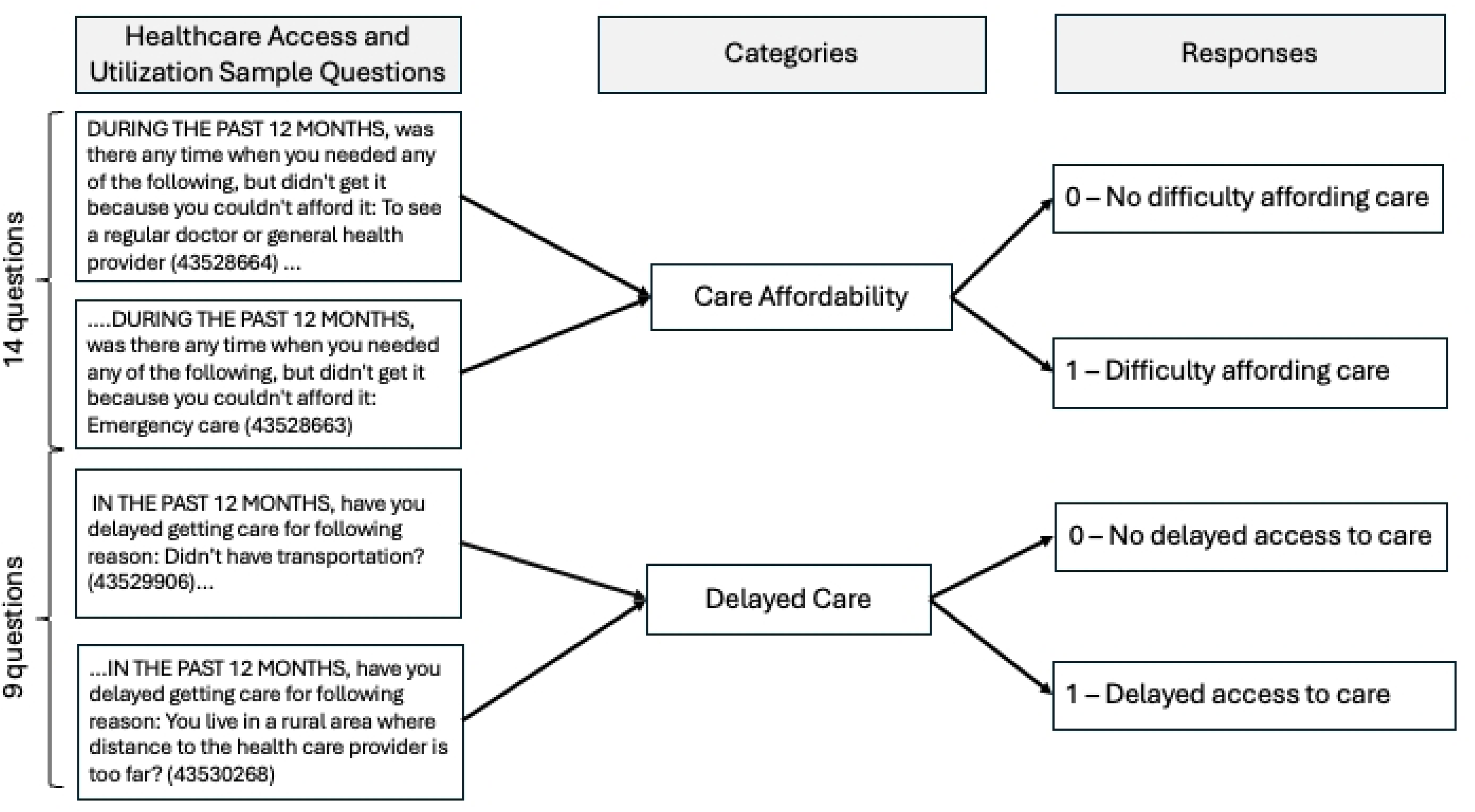
An illustration of the coding process for Healthcare Access and Utilization survey questions.

### Statistical analysis of the survey data

We analyzed the survey data on delayed care and healthcare affordability by matching the rural percentages to each *All of Us* participant’s corresponding ZIP codes and developed Empirical Cumulative Distribution Function (ECDF) plots to measure trends along the rural scale from 0% to 100% rural. The ECDF plot serves as a non-parametric method for visualizing the cumulative distribution of a dataset by displaying the cumulative probability associated with each data point. To compare and quantify statistical differences between the ECDF plots, we applied another non-parametric test, the Kolmogorov-Smirnov (KS) test. Additionally, we examined sociodemographic characteristics of participants classified in the 0% and 100% rural categories to assess differences between those residing in fully urban versus fully rural, as determined by their 3-digit ZIP code areas. These groups were selected to reflect clear, binary classifications of rurality and to minimize misclassification bias. Descriptive statistics, including frequencies and percentages for categorical variables and means with standard deviations for continuous variables, were generated for each group.

## Results

As of July 2025, the *All of Us* version 8 curated dataset had an overall sample size of 633,540 enrolled participants. We included all participants in the analysis. However, only a fraction of the total population (n = 305,860) completed the Healthcare Access and Utilization survey questions. The map in Fig 3 represents the percent rurality result of each three-digit ZIP code of *All of Us* participants in the United States.

**Fig 3.**
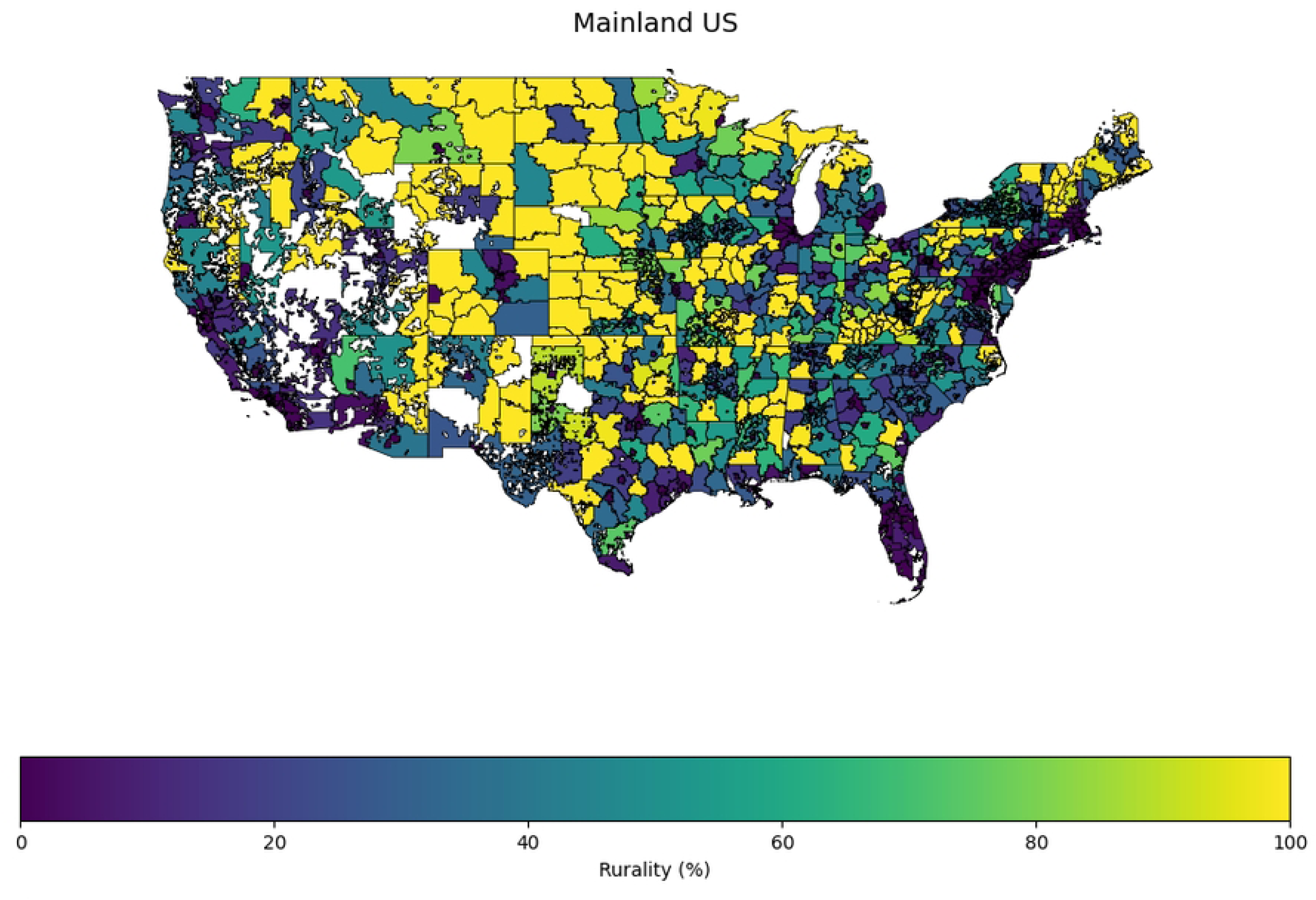
Map illustrating the three-digit ZIP codes by percent rural relating to the *All of Us* participants in the United States.

### Sociodemographic characteristics of the rural and non-rural cohort

We examined the sociodemographic characteristics of the 0% and 100% rural categories (Table 1). The 0% rural population comprised 57% of the participants (358,681 out of 633,540). Of these, 98% (354,841 out of 358,681) provided sociodemographic information. The average age was 54, with 50% identifying as White and 48% having a college or advanced degree.

**Table 1:**
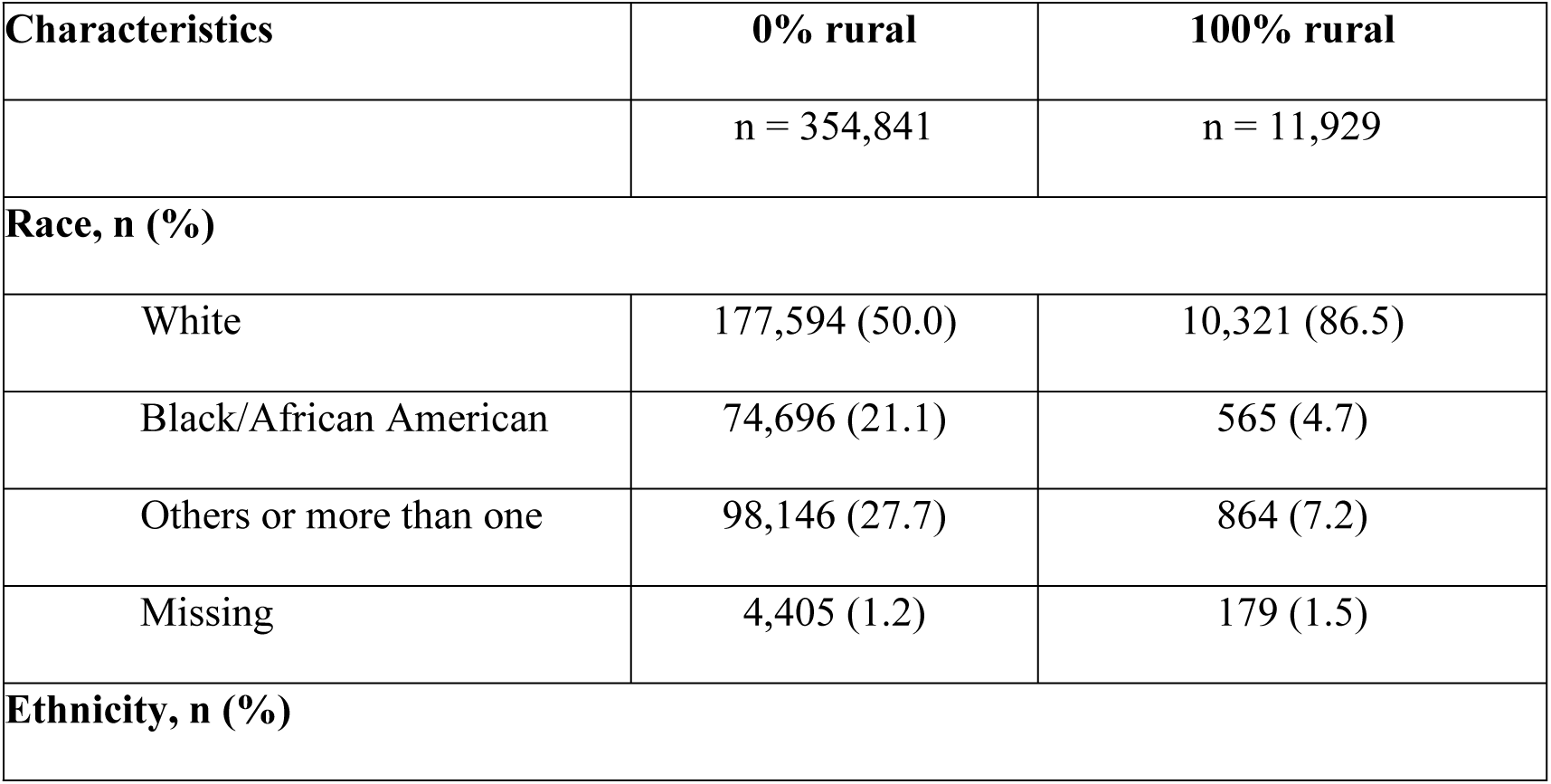
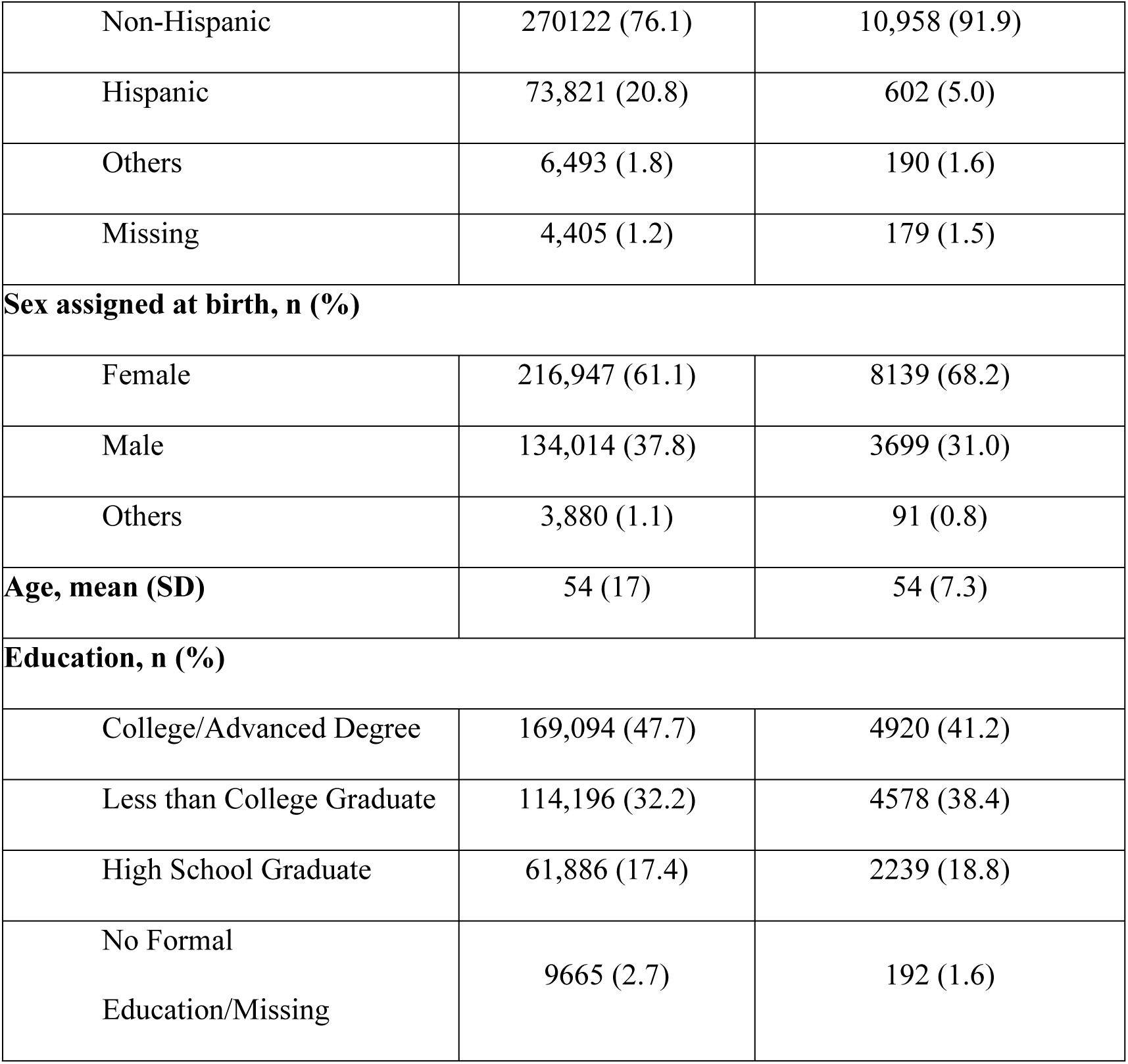
Comparison of the sociodemographic characteristics between 0% rural and 100% rural.*

| Characteristics | 0% rural | 100% rural |
| --- | --- | --- |
|  | n = 354,841 | n = 11,929 |
| **Race, n (%)** |  |  |
| White | 177,594 (50.0) | 10,321 (86.5) |
| Black/African American | 74,696 (21.1) | 565 (4.7) |
| Others or more than one | 98,146 (27.7) | 864 (7.2) |
| Missing | 4,405 (1.2) | 179 (1.5) |
| **Ethnicity, n (%)** |  |  |
| Non-Hispanic | 270122 (76.1) | 10,958 (91.9) |
| Hispanic | 73,821 (20.8) | 602 (5.0) |
| Others | 6,493 (1.8) | 190 (1.6) |
| Missing | 4,405 (1.2) | 179 (1.5) |
| **Sex assigned at birth, n (%)** |  |  |
| Female | 216,947 (61.1) | 8139 (68.2) |
| Male | 134,014 (37.8) | 3699 (31.0) |
| Others | 3,880 (1.1) | 91 (0.8) |
| **Age, mean (SD)** | 54 (17) | 54 (7.3) |
| **Education, n (%)** |  |  |
| College/Advanced Degree | 169,094 (47.7) | 4920 (41.2) |
| Less than College Graduate | 114,196 (32.2) | 4578 (38.4) |
| High School Graduate | 61,886 (17.4) | 2239 (18.8) |
| No Formal Education/Missing | 9665 (2.7) | 192 (1.6) |

The 100% rural cohort included 1.9% of enrolled participants (11,997 out of 633,540). Of these, 99% (11,929 out of 11,997) had sociodemographic information. The average age was 54, with 86% being white, 68% female, and 41% having a college education or advanced degree.

### Variations in healthcare access by geolocation

The ECDF plot (Fig 4) compares the distribution of access to care among each of the *All of Us* participants along the rural scale. A Kolmogorov-Smirnov test to compare the two distributions showed a statistically significant difference between those with and without delayed access to care (p < 0.001).

**Fig 4.**
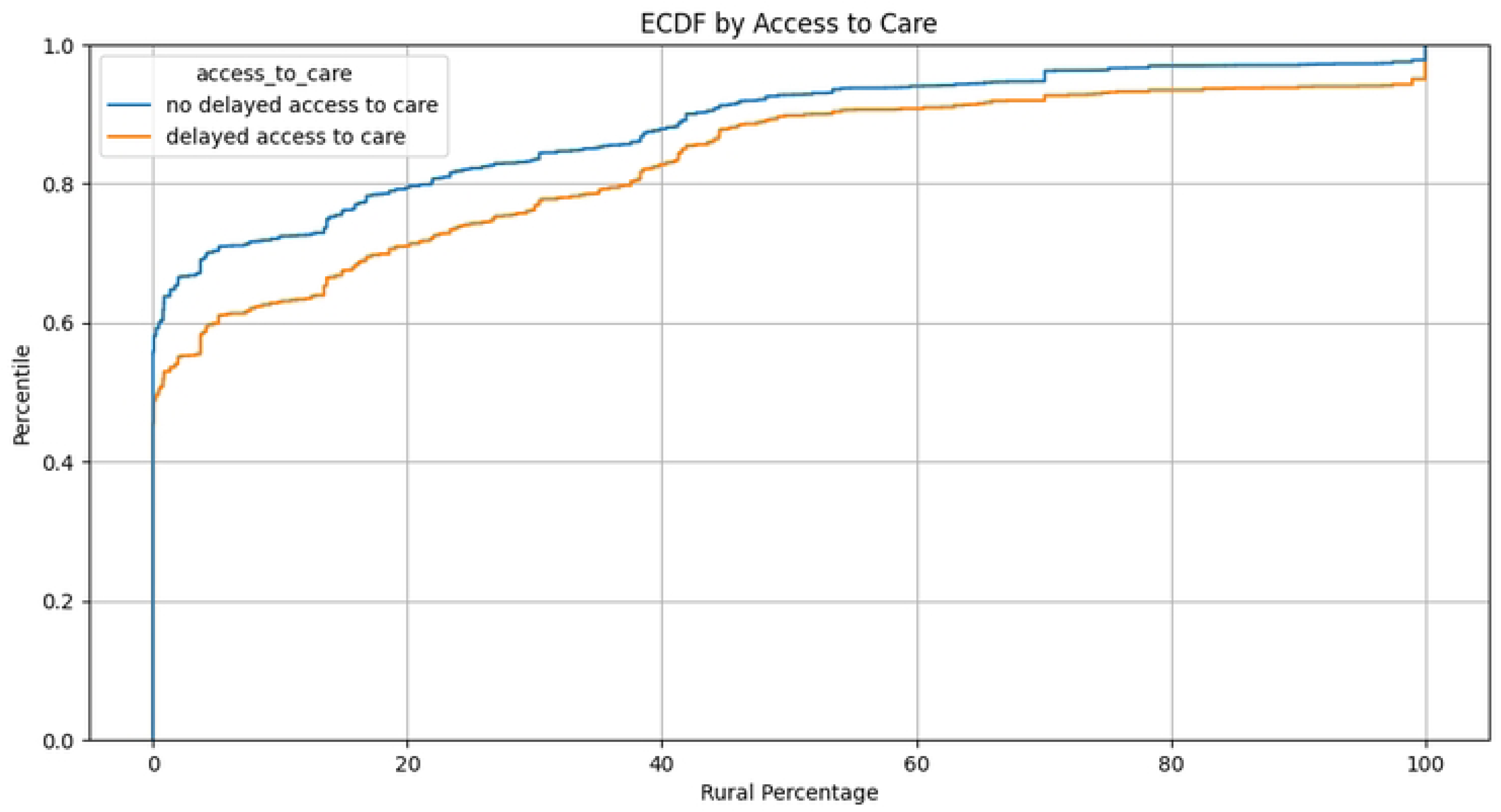
ECDF Plot showing the cumulative distribution of Access to Care along the rurality scale. At 0%, we observe that the two curves uniformly rise steeply, indicating that a significant proportion of the participants have similar levels of access to care. Subsequently, the two curves split at different percentiles, indicating a disparity in access to care with respect to the rural percentage. The orange curve (delayed access to care) indicates that as the percentile increases, those experiencing delayed access to care tend to come from areas with higher rural percentages when compared to those with no delayed access to care (blue curve).

Similarly, as shown in Fig 5, participants reporting difficulty affording care tend come from areas with slightly higher rural percentages than those with no difficulty affording care, and this difference is statistically significant (p < 0.001).

**Fig 5.**
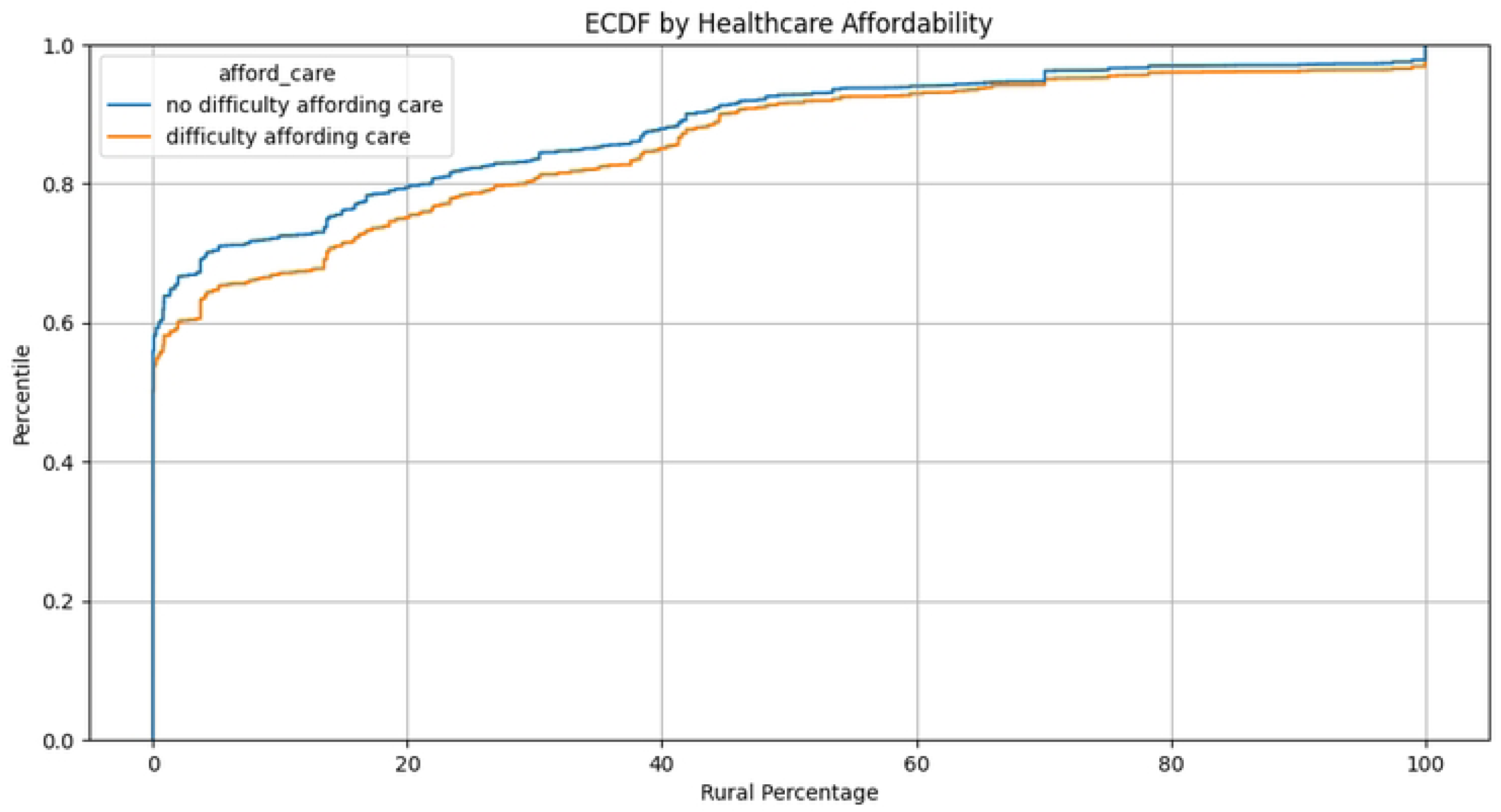
ECDF Plot showing the cumulative distribution of Healthcare Affordability along the rurality scale. Similarly, at 0%, we observe that the two curves uniformly rise steeply, indicating that a significant proportion of the participants have similar levels of healthcare affordability. Subsequently, the two curves split at different percentiles, indicating a disparity in healthcare affordability with respect to the rural percentage. The orange curve (difficulty affording care) indicates that as the percentile increases, those experiencing difficulty affording care tend to come from higher rural percentages when compared to those with no difficulty affording care (blue curve).

## Discussion

Variations in traditional definitions of rurality come from a whole host of considerations, including geographic remoteness, demographics, resources, communities’ perceptions of their own rurality, population density, commuting patterns, and proximity to large urban areas (2,4,5,7). All rural taxonomies incorporate some aspect of geography, and thus any consideration of the operationalization of “rurality” in a program, such as the *All of Us* Research Program, should start with the geographic grouping of participants in a systematic and reproducible manner.

Data from the *All of Us* research program hold significant potential for advancing the understanding of health outcomes of people living in rural America (14). The diversity of participants, combined with the breadth of data available in *All of Us*, provides a unique opportunity to explore rurality through multiple lenses, including genomics, electronic health records (EHR), and survey responses. However, to fully leverage this potential, a formal, relevant, and operational rurality scale is essential (7). A key methodological strength of our work is the application of a continuous rurality scale that is compatible with de-identified data constraints common in large, real-world health datasets. Most available resources limit geographic identifiers to 3-digit ZIP codes to protect participant privacy, making traditional, more granular rural classifications impractical. Our approach addresses this gap and offers a practical solution for researchers working within similar data environments.

A close examination of Table 1 shows the discrepancy between enrollment numbers in rural and urban locations within the *All of Us* Research Program. The number of rural participants with complete geolocation, EHR, genomic, and survey response data available is even more limited. Strengthening rural health research within *All of Us* will require both the development of a consistent rurality taxonomy and continued efforts to enhance rural participation and retention across diverse geographic regions. As an extension to our study, the three-digit ZIP code classification system (i.e., the rurality scale) can be triangulated with survey response questions, EHR data, and other information within the Researcher Workbench to define more granular cohorts for answering important rurality-related research questions. Replicating the proposed rurality scale in the *All of Us* Researcher Workbench allows for characterizing the current rural participation and the associated demographic characteristics. Of note, the inverse of this scale also presents a method to study urban populations and is a first step in realizing the abilities of geolocation function within the *All of Us* Research Program.

The *All of Us* Research Program data provides more depth than many other databases in terms of its ability to capture demographic, social, genomic, clinical, and geographic characteristics from a very large cohort (12,22). Creating ways to study rural health outcomes within the Researcher Workbench will add value to rural health research. Importantly, the program’s focus on the underserved in biomedical research could provide novel insights into the health and behaviors of the most vulnerable people living in rural areas (13,27).

## Limitations

Our study has several important limitations. The development of rural taxonomies requires consideration of as many factors of rurality as possible, including population size, remoteness, rural self-identification, commuting patterns, and proximity to urban areas. In the *All of Us* Research Program, the basis for geographic parameters is defined as the three-digit ZIP code based on the FORPH data sets. A pre-determined geographic unit is prevalent in large-scale studies and government entities because most programs are constrained by funds, data availability, and participation. As with other studies utilizing large, real-world health datasets, the methods applied in this work are driven by the requirement to protect participant privacy. Our approach was designed to meet these privacy standards while providing a practical, scalable method to characterize rurality within the constraints of available geographic information.

While some of our analyses focused on participants in the 0% and 100% rural categories, the continuous rurality scale developed here is intended for broader use. Further research is needed to establish empirically driven thresholds along the scale to differentiate urban, suburban, and rural areas, and to explore potential inflection points that may better capture gradients of rurality in relation to health outcomes.

## Conclusions

In this study, we developed a standardized approach to identify and characterize rural participation and enrollment based on the 3-digit ZIP code geolocation function within the *All of Us* Researcher Workbench. This work highlights both the potential and current gaps in understanding rural health within large-scale observational health datasets by characterizing rural participation and examining associations between rurality, delayed care, and healthcare affordability. The rurality scale proposed in this study has the potential to expand both the scope and quality of rurality-focused research within the *All of Us* Researcher Workbench. This work represents an important first step in quantifying rural representation within *All of Us* and provides the foundation for future investigations into how rurality intersects with social determinants of health and other health outcomes.

## Acknowledgments

We gratefully acknowledge *All of Us* participants for their contributions, without whom this research would not have been possible. We also thank the National Institutes of Health’s *All of Us* Research Program for making available the participant data examined in this study. This study used data from the *All of Us* Research Program’s Controlled Tier Dataset version 8, available to authorized users on the Researcher Workbench.

## Author contributions

MB conceptualized the idea. MB, TO, and VS designed the study. TO developed methods for the analysis of data. TO and VS prepared tables and figures. All authors contributed to writing and reviewing the manuscript.

## Research team positionality

Our project team includes a community scientist-clinician (MB), an informatician (VS), and an informatics trainee (TO). MB is a physician champion for the *All of Us* Research Program at the University of Arizona and Banner Health and vice chief of staff at Banner North Colorado Medical Center in Greeley, Colorado. In these roles, he advocates at the local and national levels for the ability to study rural health outcomes within the *All of Us* dataset. VS is a health systems scientist and co-lead for researcher engagement with the *All of Us* program. This project grew out of MB’s keen interest in rural health and research related to rural health outcomes.

## Data availability

The Researcher Workbench for the *All of Us* program is available at https://workbench.researchallofus.org. To gain access, researchers must complete a three-step authorization process that includes registration, completion of ethics training, and attestation to a data use agreement.

## Supporting information

**S1 Fig.** Jitter plot illustrating the distribution of three racial groups across varying rural percentages. There is a decrease in racial diversity with increasing rural percentages.

## Notes

### Competing Interest Statement

The authors have declared no competing interest.

### Funding Statement

Yes

## References

1. James CV, Moonesinghe R, Wilson-Frederick SM, Hall JE, Penman-Aguilar A, Bouye K. Racial/Ethnic Health Disparities Among Rural Adults — United States, 2012–2015. MMWR Surveill Summ. 2017 Nov 17;66(23):1–9.

2. Douthit N, Kiv S, Dwolatzky T, Biswas S. Exposing some important barriers to health care access in the rural USA. Public Health. 2015 Jun 1;129(6):611–20.

3. Hart LG, Larson EH, Lishner DM. Rural Definitions for Health Policy and Research. Am J Public Health. 2005 Jul;95(7):1149–55.

4. Long JC, Delamater PL, Holmes GM. Which Definition of Rurality Should I Use? Med Care. 2021 Oct;59(10 Suppl 5):S413–9.

5. Onega T, Weiss JE, Alford-Teaster J, Goodrich M, Eliassen MS, Kim SJ. Concordance of Rural-Urban Self-identity and ZIP Code-Derived Rural-Urban Commuting Area (RUCA) Designation. The Journal of Rural Health. 2020;36(2):274–80.

6. Danek R, Blackburn J, Greene M, Mazurenko O, Menachemi N. Measuring rurality in health services research: a scoping review. BMC Health Services Research. 2022 Nov 12;22(1):1340.

7. Bennett KJ, Borders TF, Holmes GM, Kozhimannil KB, Ziller E. What Is Rural? Challenges And Implications Of Definitions That Inadequately Encompass Rural People And Places. Health Affairs. 2019 Dec;38(12):1985–92.

8. Inagami S, Gao S, Karimi H, Shendge MM, Probst JC, Stone RA. Adapting the Index of Relative Rurality (IRR) to Estimate Rurality at the ZIP Code Level: A Rural Classification System in Health Services Research. The Journal of Rural Health. 2016;32(2):219–27.

9. Singh GK, Siahpush M. Widening Rural–Urban Disparities in Life Expectancy, U.S., 1969–2009. American Journal of Preventive Medicine. 2014 Feb 1;46(2):e19–29.

10. Differences in Rural and Urban Health Information Access and Use – Chen – 2019 – The Journal of Rural Health – Wiley Online Library [Internet]. [cited 2024 Apr 4]. Available from: 10.1111/jrh.12335

11. Probst J, Eberth JM, Crouch E. Structural Urbanism Contributes To Poorer Health Outcomes For Rural America. Health Affairs. 2019 Dec;38(12):1976–84.

12. null null. The “All of Us” Research Program. New England Journal of Medicine. 2019 Aug 15;381(7):668–76.

13. 13. All of Us Research Program | NIH [Internet]. 2020 [cited 2024 Apr 4]. All of Us Research Program | National Institutes of Health (NIH). Available from: https://allofus.nih.gov/future-health-begins-all-us

14. Ramirez AH, Sulieman L, Schlueter DJ, Halvorson A, Qian J, Ratsimbazafy F, et al. The All of Us Research Program: Data quality, utility, and diversity. Patterns (N Y). 2022 Aug 12;3(8):100570.

15. Federal Office of Rural Health Policy (FORHP) Data Files | HRSA [Internet]. [cited 2024 Mar 12]. Available from: https://www.hrsa.gov/rural-health/about-us/what-is-rural/data-files

16. How does All of Us assess diversity? What communities does All of Us consider “underrepresented in biomedical research?” – All of Us Research Hub [Internet]. [cited 2024 Apr 4]. Available from: https://www.researchallofus.org/faq/how-does-all-of-us-assess-diversity-what-communities-does-all-of-us-consider-underrepresented-in-biomedical-research/

17. Le Lait MC, Martinez EM, Severtson SG, Lavery SA, Bucher-Bartelson B, Dart RC. Assessment of prescription opioid intentional exposures across the rural-urban continuum in the United States using both population and drug availability rates. Pharmacoepidemiology and Drug Safety. 2014;23(12):1334–7.

18. National Rural Health [Internet]. [cited 2024 Sep 23]. All of Us Research Program aims to enroll 1 million participants | NRHA. Available from: https://www.ruralhealth.us/blogs/2018/09/all-of-us-research-program-aims-to-enroll-1-million-participants

19. All of Us Research Program | NIH [Internet]. 2020 [cited 2024 Sep 23]. The All of Us Journey Brings More Research Program Features to All of You. Available from: https://allofus.nih.gov/news-events/announcements/all-us-journey-brings-more-research-program-features-all-you

20. Data Standardization – OHDSI [Internet]. [cited 2025 Apr 23]. Available from: https://www.ohdsi.org/data-standardization/

21. Data Methods – All of Us Research Hub [Internet]. [cited 2024 Mar 15]. Available from: https://www.researchallofus.org/data-tools/methods/

22. User Support [Internet]. 2023 [cited 2024 Apr 4]. Accessing geolocation data. Available from: https://support.researchallofus.org/hc/en-us/articles/4583333207956-Accessing-geolocation-data

23. Rights (OCR) O for C. Guidance Regarding Methods for De-identification of Protected Health Information in Accordance with the Health Insurance Portability and Accountability Act (HIPAA) Privacy Rule [Internet]. 2012 [cited 2024 Aug 19]. Available from: https://www.hhs.gov/hipaa/for-professionals/privacy/special-topics/de-identification/index.html

24. USA ZIP Code Areas – Overview [Internet]. [cited 2024 Mar 12]. Available from: https://www.arcgis.com/home/item.html?id=8d2012a2016e484dafaac0451f9aea24

25. Survey Explorer – All of Us Research Hub [Internet]. [cited 2024 Mar 12]. Available from: https://www.researchallofus.org/data-tools/survey-explorer/

26. All of Us Public Data Browser [Internet]. [cited 2025 Apr 1]. Available from: https://databrowser.researchallofus.org/survey/health-care-access-and-utilization

27. Chandler PD, Clark CR, Zhou G, Noel NL, Achilike C, Mendez L, et al. Hypertension prevalence in the All of Us Research Program among groups traditionally underrepresented in medical research. Sci Rep. 2021 Jun 22;11(1):12849.

